# Detection of early-stage cancer using similarity-based classification of cell-free DNA fragmentation patterns

**DOI:** 10.64898/2026.11.08.26366892

**Authors:** Derrick E. Wood, Bari J. Ballew, Joseph Roy

## Abstract

Current cancer screening efforts face the dual problems of low patient adherence and a lack of available tests for many cancers, which motivates the development of less invasive multi-cancer early detection strategies. Such strategies will likely involve the use of liquid biopsies, which can analyze biomarkers found in blood for the presence of cancer. We introduce Blackjack, a new detection approach that focuses on the fragmentation patterns of plasma-derived cell-free DNA (cfDNA). Blackjack characterizes a person’s cfDNA fragmentation sites with a position probability matrix (PPM) that encodes information about the sequence context of the sites. For classification, Blackjack computes two features that represent the similarity of an individual’s PPM to those of known healthy and cancer cohorts. With these two features, Blackjack then uses logistic regression to classify a new sample as being from a healthy individual or a person with cancer. In cross-validation experiments using two external whole genome sequencing datasets, Blackjack showed high performance, achieving a minimum area under the receiver operating characteristic curve (AUC) of 0.993 and minimum sensitivity (Sn) of 91.8% (at 98% specificity). This high performance was also present for stage I (AUC 0.991, Sn 89.8%) and stage II cancers (AUC 0.993, Sn 90.2%), and across eight different cancers (minimum AUC 0.985, minimum Sn 80.0%). We demonstrate that Blackjack’s accuracy is stable at low simulated sequencing depths (∼0.24x), and that tissue of origin prediction using Blackjack’s similarity measures is high (minimum 82% accuracy of the top two predicted tissues). Our findings show that cfDNA fragmentation site contexts have valuable signals for early cancer detection, and that developing diagnostics with machine learning can benefit from a focus on having a small number of informative features.

## Introduction

Detection of cancer in early stages is known to improve survival and the efficacy of treatment [1,2]. To that end, regular screening tests for certain cancers are recommended [3–5]. These tests often require scheduling appointments [6], can require uncomfortable preparation [7], and have procedures that can be painful for the patient [8]. All of these factors have been implicated in low patient adherence rates for screening. Further hampering the goal to lower cancer mortality is a lack of recommended screening tests for most solid tumors. As of 2026, the United States Preventive Services Task Force only recommends widespread screening for breast, colorectal, lung, and cervical cancers [9–12]. Over a million new cases from other cancers are estimated to occur in 2026 in the United States alone [2], highlighting the impact of this screening gap.

A possible solution to the problem of low adherence has emerged in recent years with the development of blood-based cancer tests; these are referred to by some as “liquid biopsies” [13]. These tests, which only require a patient to arrive without an appointment at a laboratory for a routine blood draw, may increase adherence for screening by removing some of the factors discussed earlier. Outside of screening, liquid biopsy tests have also been used for tasks such as tumor profiling [14] and detection of residual or recurrent cancer after treatment [15]. While protein biomarkers such as CA125 have been known to be of value in cancer screening for decades [16–18], the field has recently seen the development of blood-based cancer screening tests that look at cell-free DNA (cfDNA). These cfDNA-focused tests examine biomarkers such as methylation [19], somatic mutations [20], and aneuploidy [21]. Such tests have been part of the recent industry growth around multi-cancer early detection [22], which seeks to detect multiple cancers with a single test, and may serve to detect cancers for which screening tests do not presently exist.

Within the field of liquid biopsy, some tests focus on the fragmentation pattern of cfDNA to detect cancer; this is often termed “fragmentomics” [23]. Fragmentomics typically examines whole genome sequencing (WGS) of cfDNA to determine an individual’s fragmentation pattern. These patterns reflect various cell types of the cfDNA’s origin, and healthy people tend to have cfDNA that is primarily from myeloid and lymphoid cells [24]. Healthy individuals also have similar patterns to each other [25]. Circulating tumor DNA (ctDNA) will distort an individual’s pattern by contributing DNA from a cell type that is different (or is in different proportions) from what is seen in healthy individuals. In general, fragmentomics is seeking to detect this distortion to diagnose cancer.

Key questions in a fragmentomics approach are how a pattern is represented, and how various individuals’ representations are compared to each other. Patterns are often represented using information about the cfDNA fragment length distribution [26,27], the sequence context around the cfDNA fragment sites [28], or statistics relating to the frequency of observing fragment ends at certain genomic loci [29]. From a computational perspective, some of these representations can be viewed as a high-dimensional vector. Some fragmentomics approaches use dimensionality reduction techniques to project these vectors into a lower-dimensional space [30]; these spaces can allow for classification models that would be less likely to overfit to training data. Other approaches may forgo dimensionality reduction and build machine learning (ML) classification models that use the elements of the high-dimensional vector as features [31].

We introduce here our fragmentomics approach, named Blackjack. Blackjack represents an individual’s cfDNA fragmentation pattern using a dinucleotide position probability matrix [32] to encode sequence context information about the sites of fragmentation. We then use distance and similarity transformations as a dimensionality reduction technique, directly taking advantage of the property that healthy peoples’ fragmentation patterns are similar. In a two-dimensional space defined by similarity to the two sample classes (healthy and cancer), we show that healthy and cancer samples are highly distinguishable in two separate datasets.

## Materials & Methods

### Creating an individual’s position probability matrix

To create a position probability matrix (PPM) for an individual, we began from the FinaleDB [33] data that indicated the mapping location for each mate pair of sequencing reads. We used the hg38 (GRCh38) reference human genome to extract a 6 nucleotide sequence that was centered on each fragment end. Each fragment contributed two hexamer sequences, and we reverse complemented the hexamer centered on the rightmost (i.e., genomic 3’) end of a fragment. We rejected a fragment’s pair of hexamers if any of the following were true: (a) the fragment did not align to an autosomal chromosome, (b) the fragment’s minimum mapping quality (MAPQ) score is less than 60, or (c) either of the fragment’s hexamer contexts in the genome contain a non-ACGT nucleotide code. The hexamers that remain were used to create a 16 x 5 matrix of dinucleotide-position counts, where a cell indicates how many times a dinucleotide was seen at a given position. The count matrix was then normalized so that each column sums to 1 to create a PPM, as seen in Fig. 1A. Due to the positional independence assumption used by PPMs [34] and the unit-sum constraint of each column of the PPM, only 15 free parameters exist within each column, and a single hexamer contributes to a frequency in each of the five columns. Thus, our PPMs have a total of 80 elements and 75 free parameters.

**Figure 1.**
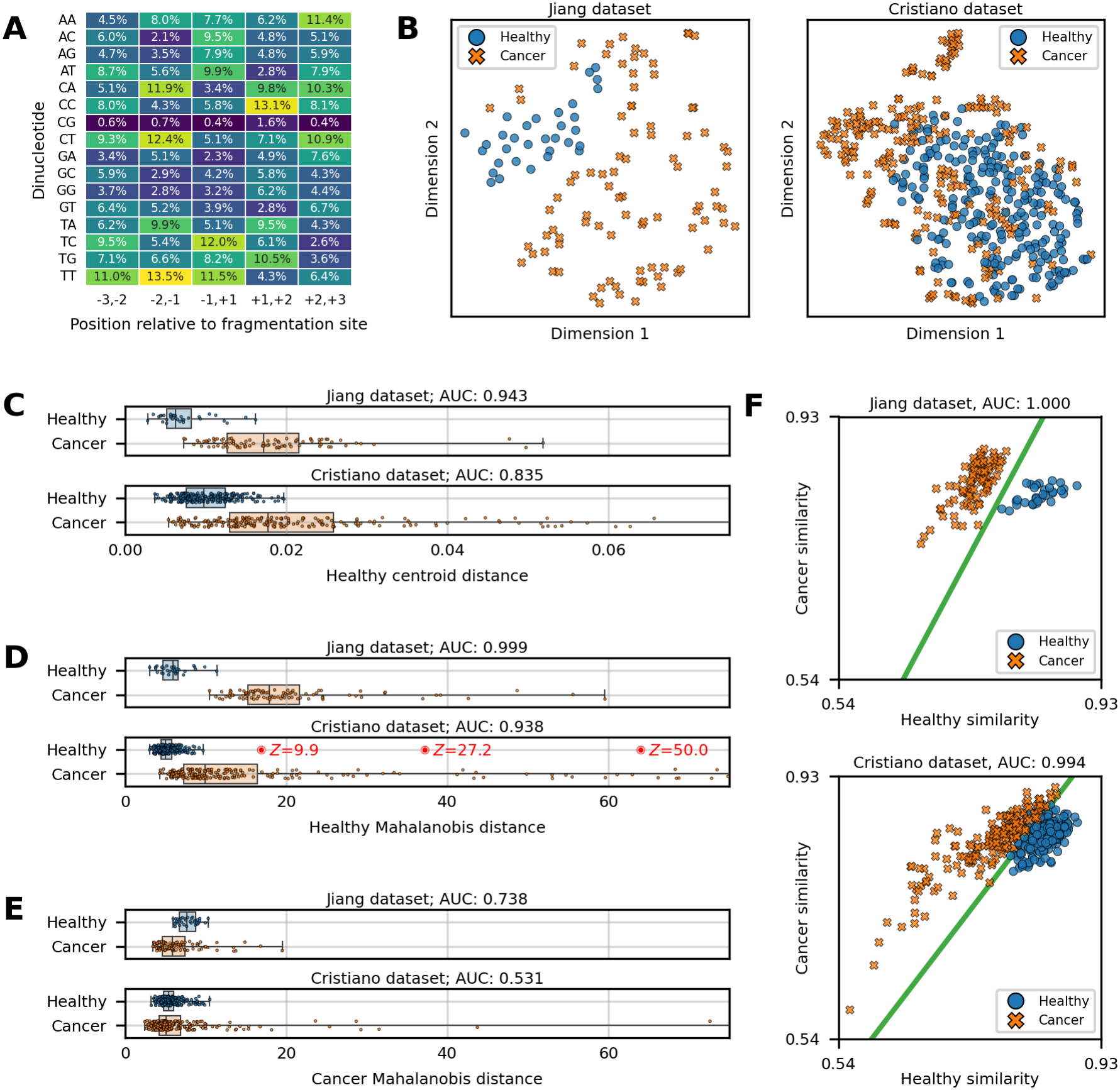
Elements of our similarity-based classification method. **(A)** A dinucleotide position probability matrix (PPM), indicating the percentage of fragments that had a given dinucleotide at a position relative to the fragmentation site. We use a PPM to represent a person’s cfDNA fragmentation pattern. **(B)** A t-distributed stochastic neighbor embedding (t-SNE) visualization of our two datasets, where each point is a single individual’s PPM. **(C)** Healthy centroid distance (HCD) values for the two datasets, with 12 high distance (> 0.075) cancer points not displayed. **(D)** Healthy Mahalanobis distance (HMD) values for the two datasets, with 5 high distance (> 75) cancer points not displayed. Three removed outliers from the Cristiano dataset are highlighted here with red dots and *Z* scores based on the HMD values of the remaining healthy samples. **(E)** Cancer Mahalanobis distance (CMD) values for the two datasets, with 1 high distance (> 75) cancer point not displayed. **(F)** Class similarity plots for the two datasets. In each case, a green line shows the boundary learned by a logistic regression (LR) classifier in the two-dimensional similarity space. The area under the curve (AUC) is reported using receiver operating characteristic analysis for the HCD, HMD, CMD, and similarity LR metrics.

### External data collection

We downloaded fragment alignment data from FinaleDB that corresponded to WGS data from two different publications. We specifically chose to work with the data from what we term the “Jiang dataset” [35] and the “Cristiano dataset” [25] because they had samples from both healthy donors and individuals with cancer. This was necessary for the purposes of developing a tool to distinguish between individuals who were healthy and those with cancer. We gathered metadata for the samples from both the FinaleDB webpages as well as the supplementary data of the Cristiano publication [25].

The Jiang dataset contained data from four categories of samples: healthy, cirrhosis, hepatitis B, and liver cancer. We focused this work on the 32 healthy and 90 liver cancer samples in that dataset. 85 of the dataset’s cancer samples were from stage A liver cancer, and 5 were from stage B liver cancer; we did not have any indication as to which samples were stage A or B.

The Cristiano dataset had data from eight different types of cancer, as well as healthy samples. In working with this dataset, we had to exclude many samples from our analyses for various reasons. Some samples were from technical replicates of other samples; we retained only one replicate per donor for our analyses so that a single individual donor would not overly influence the results. So that our analyses of per-stage and per-cancer accuracy (Fig. 2C) could have sufficient samples, we excluded cancer samples that did not have at least 9 other samples of the same stage or cancer type; this removed the sole duodenal cancer sample, two stage 0 samples, and one stage X sample. We also removed from our analyses any samples not listed as being “preoperative treatment naïve,” which removed two non-cfDNA samples as well as all stage IV lung cancer samples (collected at multiple timepoints per patient) that were from a set of 19 patients that were “undergoing treatment with anti-EGFR or anti-ERBB2 therapy” [25]. We did not use two samples in FinaleDB relating to one pancreatic cancer patient (CGPLPA95) because we did not have two entries in the Cristiano paper’s supplementary data for that patient. Finally, we removed three healthy samples for being strong outliers, as we describe later in this section. This left this dataset with 241 healthy samples and 211 cancer samples. The distribution of the 211 cancer samples by stage and cancer type is shown in Supplementary Fig. S1.

**Figure 2.**
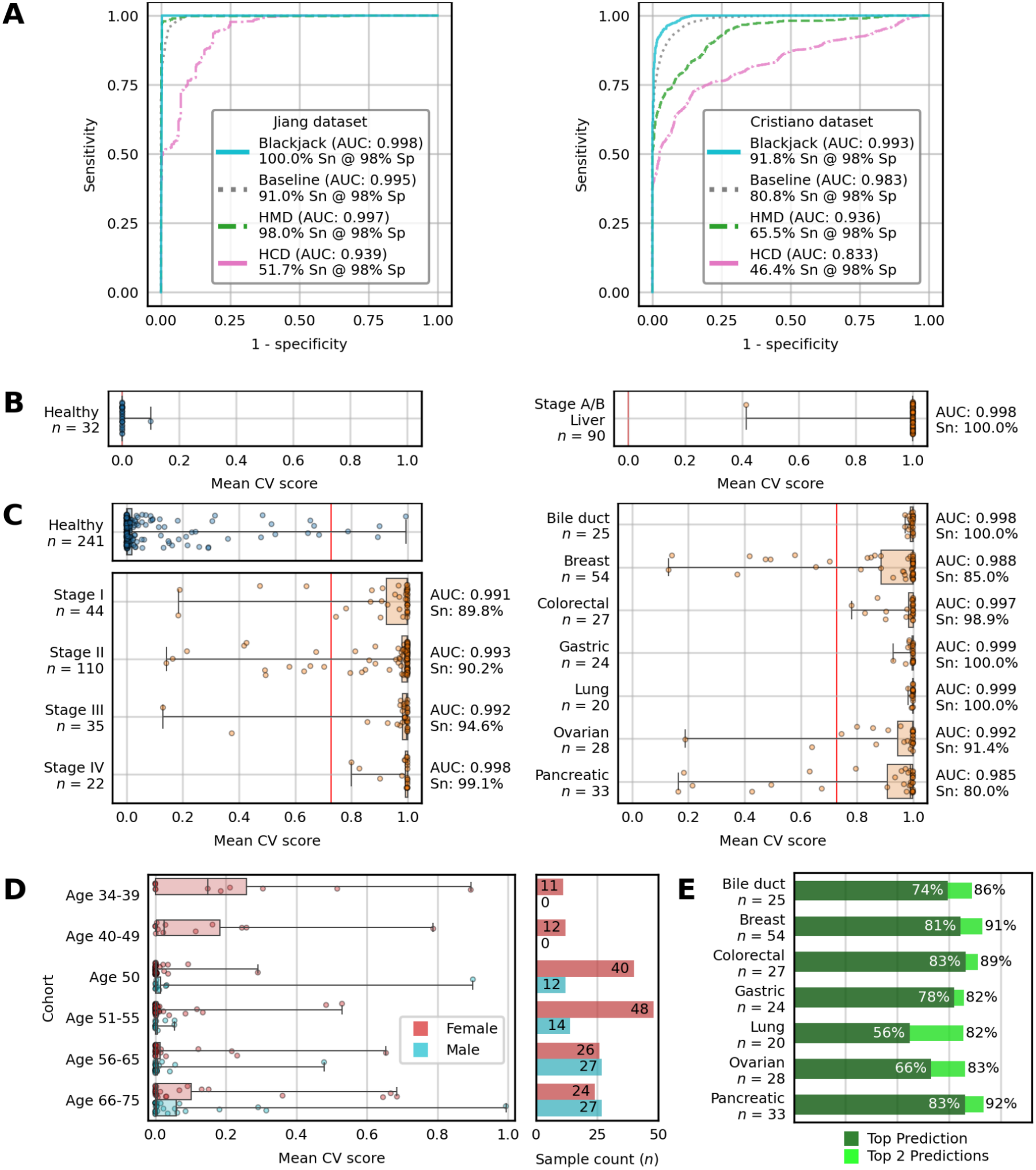
Cross-validation results. **(A)** Receiver operating characteristic curves for the four prediction approaches we evaluated in 10-fold cross-validation (CV): Blackjack; a baseline approach using an Extra-Trees model and principal component analysis; healthy Mahalanobis distance (HMD); and healthy centroid distance (HCD). Area under the curve (AUC) and sensitivity (Sn) at 98% specificity (Sp) are shown for each approach. **(B)** Mean CV score distributions for the Blackjack approach on the Jiang dataset. AUC and Sn at 98% Sp are shown for cancer samples, and the 98% Sp score threshold is shown as a vertical red line. **(C)** Mean CV score distributions for the Blackjack approach on the Cristiano dataset, stratified by healthy/cancer status, cancer stage, and cancer type. **(D)** Mean CV score distributions for the Blackjack approach on the healthy samples of the Cristiano dataset, stratified by age and gender. **(E)** Tissue of origin prediction accuracy. For each cancer type, accuracy of the top prediction and accuracy of one of the top two predictions is shown.

### Calculation of distance and similarity

During model training, and for all the distance and similarity methods shown in Fig. 1, when computing the healthy centroid distance or healthy Mahalanobis distance for a healthy PPM *p*, we calculated the centroid (and covariance matrix, for the Mahalanobis distance) of the training data’s healthy cohort with *p* excluded from the calculation. We did this to ensure that the estimators of the healthy distribution (the centroid and covariance matrix) were not skewed toward the single point *p* that we were also examining. A similar leave-one-out fitting operation was done when computing cancer Mahalanobis distances to a cancer PPM.

In computing the Mahalanobis distance, our software library (scipy) required an inverse covariance matrix. We added a small constant to each diagonal element of the covariance matrix prior to inversion so that any collinearity between elements of PPMs would not inhibit inversion; this is a technique known as zeroth-order Tikhonov regularization [36]. To compute a class Mahalanobis similarity value, we raised the healthy (or cancer) Mahalanobis distance to a small constant negative exponent. In Fig. 1, we used 10^-7^ as the constant for the diagonal elements, and -0.1 as the constant exponent. In our cross-validation experiments for cancer detection shown in Fig. 2, our Blackjack approach uses a grid search within each training fold to select the best performing combination (by AUC) of diagonal constant and constant exponent, so that we are not affecting the classifier’s performance by making the decision ahead of time.

### Outlier removal using healthy Mahalanobis distance

To detect outliers among the healthy cohort in each dataset, we used a method suggested previously [37]. We began by computing the Mahalanobis distance (MD) to the other healthy samples of a dataset. We then performed a Chi-squared test on the square of the MD, using 75 degrees of freedom (due to the 75 free parameters of our PPMs). Samples with a *p*-value less than 10^-6^ were considered to be outliers; only the Cristiano dataset had any outliers detected.

As a separate check to ensure the outliers were indeed strongly dissimilar to the other healthy samples, we re-computed the MD for each of the 241 remaining healthy samples in the Cristiano dataset to the other 240 healthy samples. Calculating the mean and standard deviation of these MD values allowed us to obtain *Z* scores, view the distribution of *Z* scores among the 241 inlier healthy samples, and verify that the 3 outliers had extremely high *Z* scores.

### Cross-validation experiments

Within each round of our *k*-fold cross-validation (CV) experiments, a dataset was randomly divided into *k* stratified folds, where each fold had approximately equal proportions of healthy samples and equal proportions of each type of cancer (e.g., breast cancer, lung cancer). We performed 10 rounds of CV for each experiment, and each round tested on each fold once. This meant that within our 10-fold CV experiments, each individual point within our datasets was used for training 90 times and testing 10 times. We used 10-fold CV for both our cancer detection and tissue of origin prediction experiments (Fig. 2), and a 2-fold CV experiment to test robustness of our cancer detection approaches (Supplementary Fig. S2).

We tested three different distance- or similarity-based classification approaches in our cancer detection CV experiments. The first two model the healthy distribution with either a centroid (“HCD”) or a centroid and covariance matrix (“HMD”) and use logistic regression (LR) to treat distance to that distribution as a single classification feature. The third (“Blackjack”) models both healthy and cancer distributions with centroids and covariance matrices and uses similarity to both distributions as inputs to a two-feature LR model.

Regularization penalties can be employed with LR in an effort to keep coefficient magnitudes low and avoid overfitting [38]. As described earlier, we do use a form of regularization for the covariance matrices when computing Mahalanobis distances. However, our three LR models used for cancer detection had a maximum of two features, and were unlikely to be overfit with the sizes of the training sets in our CV experiments. For this reason, we did not use any additional regularization penalties when fitting the LR classifiers.

We additionally tested a baseline classification approach that used an extremely randomized trees (Extra-Trees) [39] model along with principal component analysis (PCA). The PCA transformation of the 80 PPM features resulted in features for the Extra-Trees model that were far less correlated. We selected the Extra-Trees model as our baseline model because of its speed of model fitting and its robustness to overfitting as compared to other tree-based ensemble models, such as a random forest.

The confidence intervals for the reported statistics in our cross-validation experiments (Supplementary Tables S1 and S2) were determined by 1000 bootstrap resamplings of the patients relevant to each statistic.

### Software and data availability

We used the scikit-learn package [40] (v0.23.2) to build our ML models and to calculate classification statistics; matplotlib [41] (v3.5.1) and seaborn [42] (v0.11.2) to create plots; pandas [43] (v1.3.5) and numpy [44] (v1.21.5) for operating on tabular data and various arithmetic operations; and scipy [45] (v1.8.0) for statistical tests and distance calculations. For extraction of hexamers from the reference genome, we used bedtools [46] (v2.30.0) and samtools [47] (v1.13).

The software we wrote to extract hexamers and perform the work shown in this article is freely available to use and to download in a data archive from Zenodo [48].

## Results

### Representation of an individual’s fragmentation pattern

Our implementation of fragmentomics focuses on the sequence contexts around the sites of cfDNA fragmentation. Sequence contexts around cfDNA fragment ends are similar in fragments from the same organ, and different in fragments from different organs [28]. This property of the sequence contexts has been associated with differences in nuclease activity between cell types [23], with the DNA endonuclease DNASE1L3 thought to be directly associated with at least some of the differences in sequence contexts [28]. We sought to use this property in the development of Blackjack.

Working from paired-end whole genome sequencing (WGS) data, we examine the 6 bp (hexamer) window centered on the observed fragmentation site, treating the first base of each read of a mate pair as a fragment end. In this way, each fragment of cfDNA provides us two data points, with a hexamer centered on each end of the fragment. We use the many hexamer contexts collected from an individual to create a position probability matrix (PPM) that indicates the frequency at which a given dinucleotide is seen at each position (relative to the fragmentation site). Each column of a PPM is an empirical probability mass function (PMF) for the specific position represented by that column, and so the values within a column must sum to one. We selected the PPM as the representation for an individual’s cfDNA fragmentation pattern in part due to its low complexity. Representing a collection of hexamer contexts with a dinucleotide PPM only requires 80 quantities to be estimated (16 dinucleotides x 5 positions within the hexamer).

In the example PPM shown in Fig. 1A (a healthy sample selected at random), the PPM indicates that 13.1% of fragment ends began with two consecutive C bases, and that 12.4% of fragment sites had a CT dinucleotide immediately upstream. We used a dinucleotide PPM instead of the more traditional mononucleotide PPM so that we could capture the information that exists between adjacent nucleotides; similar decisions have been made by others with position weight matrices [49], which are close relatives of PPMs. As an example, in all five positions of the PPM in Fig. 1A, the CG dinucleotide is present in a low percentage of fragments (maximum frequency of 1.6%). This reflects the low prevalence of CpG sites in the genome [50] and is a more accurate representation of the CG dinucleotide’s prevalence than would be implied by a mononucleotide PPM. The dinucleotide PPM is also similar to transcription factor flexible models [51], used to describe transcription factor binding site motifs.

Using a PPM to represent a single individual as an 80-dimensional point also enables us to compare two individuals by computing the distance between two points. We have found various distance metrics to be of similar value in our investigations, but in our work that specifically compares one PPM to another (Figs. 1B and 1C), we use the Jensen-Shannon distance [52] due to its general suitability for use when comparing PMFs.

### External datasets used for exploration

In developing our methods, we used two sets of WGS data. These were introduced in prior work by others, and we refer to the datasets by the names of those papers’ first authors. The Jiang dataset [35] contains both healthy samples and early-stage liver cancer samples. The Cristiano dataset [25] contains healthy samples and cancer samples across all stages and seven different types of cancer. Rather than work with the raw sequencing data itself, we obtained the fragmentation site information from FinaleDB [33]. FinaleDB obtained the raw sequencing data from several projects, preprocessed and aligned that data, and now publicly provides information on the alignment position of the reads without revealing mutational data that could identify the sample donors.

### Distance to healthy samples is a valuable biomarker

A common step in visualization of high-dimensional data is to reduce the dimensionality using t-distributed stochastic neighbor embedding [53] (t-SNE). A t-SNE visualization of a dataset projects the high-dimensional data into a low-dimensional space, attempting to keep points that were close together in the original space near to each other in the new space. Viewing a t-SNE representation of the Jiang dataset, we saw that the healthy samples tended to be closer to each other than they were to cancer samples (Fig. 1B). We saw a similar although less pronounced form of this in the Cristiano dataset. This led us to hypothesize that comparing a novel sample to the distribution of known healthy samples would provide valuable information as to the sample’s cancer status.

One way to compare a point *p* against a set of other points is to compute the centroid of the set, and then measure the distance between *p* and the centroid. We see in Fig. 1C that the healthy centroid distance (HCD) does distinguish healthy from cancer samples well in both datasets. When computing the receiver operating characteristic (ROC) curve [54], the area under the curve (AUC) is 0.943 for the Jiang dataset and 0.835 for the Cristiano dataset. The HCD is a metric that relies on only one estimator (the centroid), and so is likely to avoid overfitting, although that simplicity may be insufficient for describing the distribution of healthy samples in the 80-dimensional space of our PPMs.

To better model the distribution of healthy samples, we can also use a covariance matrix of the healthy PPMs’ features, instead of just the centroid. The Mahalanobis distance [55] allows us to compare a point to a set of points, using the set’s centroid and covariance matrix. We can also compute the healthy Mahalanobis distance (HMD) analogously to the HCD. The HMD outperforms the HCD in separating healthy samples from cancers, with an AUC of 0.999 for the Jiang dataset and 0.938 for the Cristiano dataset (Fig. 1D).

The HMD also allows us to identify outliers in the healthy cohort. We used the HMD to identify 3 outlier samples out of 244 samples within the Cristiano dataset that were labelled as healthy (Fig. 1D). Using the remaining 241 healthy samples, we can recompute the HMD for those samples, and calculate *Z* scores for those 241 samples by subtracting the mean HMD and then dividing by the standard deviation of the HMD values; those *Z* scores range from -1.9 to 3.8. In contrast, the outlier *Z* scores are 9.9, 27.2, and 50.0. Without the ability to follow up with individual donors, it is impossible for us to conclude what caused a sample to be an outlier. However, undiagnosed cancers are possible in patients otherwise believed to be healthy. These outlier samples constitute 1.2% of the dataset’s healthy cohort. This rate of outliers is similar to that of the discovered cancer rate of 0.97% reported in a previous clinical study of a population of women aged 65-75 [20]. Regardless of the cause underlying the outlier nature of these 3 samples, we considered them to be inappropriate for use in our ML work, and removed them from our other analyses. All statistics we report are conditioned on the removal of these 3 outliers.

### Class similarity-based classification

Although the HMD appears to be a valuable biomarker in its own right, we were interested in obtaining further separation between healthy and cancer samples. To do so, we employed two additional ideas: the calculation of a cancer Mahalanobis distance (CMD), and conversion of the two MDs into similarity measures.

Much as we can compute the distance from a point to the set of healthy samples, we can also compute the distance from a point to the set of cancer samples. Our t-SNE visualizations (Fig. 1B) suggest that the cancer samples lack the same intraclass similarity that is found in the class of healthy samples. Indeed, we find that the CMD is a much weaker biomarker than the HMD, with an AUC of 0.738 in the Jiang dataset, and 0.531 in the Cristiano dataset (Fig. 1E). However, the CMD and HMD are not highly correlated; the two distances have Spearman correlation coefficients of 0.12 in the Jiang dataset and 0.38 in the Cristiano dataset. This orthogonality suggests that they may be valuable for discriminating between classes when used in combination with each other.

When visualizing the data in a two-dimensional space with these two features, the high distances of some samples can make seeing a potential boundary between the two classes difficult. While distance functions must fulfill certain mathematical properties, if we are willing to relax some of those properties, we can transform these distances into similarity measures [56]. Such similarity measures can maintain the relative order of the points along an axis, and preserve much of the informative value of the distance metric while making both visualization and classification easier. We computed similarity measures here by raising the two class MDs to an exponent. In this two-dimensional similarity feature space, we see that the two datasets can be classified well using a traditional logistic regression (LR) model (Fig. 1F). Using LR models, the two-dimensional class Mahalanobis similarity space achieves an AUC of 1 for the Jiang dataset, and 0.994 for the Cristiano dataset. Our Blackjack classification approach uses LR models in this space to distinguish between healthy and cancer samples.

### Cross-validation shows high performance in two datasets

The previous mentions of AUC statistics are all based on retrospective analysis. To better approximate the performance of a method on novel data, we performed cross-validation (CV) experiments (Fig. 2A, Supplementary Table S1). In CV, a portion of a dataset is withheld from model training, and then the model is tested on the withheld data; this process is repeated multiple times with different subsets of the data withheld for testing. Our CV experiments tested the HCD, HMD, and Blackjack approaches, using LR to classify healthy and cancer samples with each approach. We also compared against a baseline approach that used principal component analysis on the PPMs before using an Extra-Trees model for classification.

Because cancer is expected to have low prevalence in a screening population, screening tests are expected to have high specificity to maintain an appropriate positive predictive value [57]. For this reason, we evaluated our CV experiments by measuring not only AUC but also sensitivity at a 98% specificity threshold. On the two datasets, the Blackjack approach achieved a minimum AUC of 0.993, and sensitivities of 100.0% (Jiang dataset) and 91.8% (Cristiano dataset). The Blackjack approach also met or exceeded the performance of the other distance-based approaches in terms of AUC and sensitivity in both datasets. Our baseline approach, designed to use our PPM frequencies in a more traditional ML pipeline, underperformed Blackjack in both datasets, although it did outperform the HMD approach on the Cristiano dataset and was similar to the HMD approach’s results on the Jiang dataset. These results compared to the baseline approach suggest Blackjack’s focus on similarity provides predictive utility beyond that provided by the PPM features themselves.

While the Jiang dataset’s cancer samples are mostly stage A liver cancer (Fig. 2B), the multiple cancer types and stages present in the Cristiano dataset allow us to examine performance of the Blackjack approach across various strata of interest (Fig. 2C, Supplementary Table S1). As may be expected given the higher amounts of ctDNA present in later stage disease [58], AUC and sensitivity are higher in stage IV cancers than in earlier-stage cancers. Nonetheless, Blackjack’s accuracy in early stages is still high, with a stage I AUC of 0.991 and stage I sensitivity of 89.8%. Performance of Blackjack across the various cancer types is high, with the lowest AUC and sensitivity found in the pancreatic cancer subset (0.985 and 80.0%, respectively). Our Blackjack approach sees sensitivities of over 99% for bile duct, gastric, and lung cancers. We report AUC and sensitivity results for each stage within each type of cancer of the Cristiano dataset in Supplementary Fig. S1.

The above CV results were all generated via a set of 10-fold CV experiments. We also wanted to investigate the robustness of these methods by using 2-fold CV experiments (Supplementary Fig. S2). Such experiments would have substantially less training data available to the methods (training on only 16 healthy and 45 cancer samples in the Jiang dataset) and so would provide a strenuous test to the methods. As expected, the 2-fold CV AUC and sensitivity statistics were less than each methods’ corresponding statistic in the 10-fold CV experiments. The minimum AUC across the two datasets for Blackjack in these 2-fold CV experiments was 0.989, and the minimum sensitivity was 86.5%. Of particular note is the performance in these 2-fold CV experiments of the HCD approach, which still achieves a minimum AUC of 0.834 and minimum sensitivity of 47.1%, while relying on only one feature (the distance to the healthy centroid).

### Putative presence of fetal cfDNA detected as aberrance

In examining the healthy cohort within the Cristiano dataset, we noticed that none of the 23 samples with an age less than 50 years old (y.o.) were from a male donor. We divided this healthy cohort into age groups of roughly equal size (but of no more than 10 years in range), and then examined the mean Blackjack CV scores per sample (Fig. 2D). The samples that were from donors < 50 y.o. have higher scores than any of the older age groups, when evaluated by the median sample’s mean score. Higher scores should normally correspond to a higher likelihood of cancer. In examining samples labelled as “healthy”, we might expect the older age groups to have a slightly elevated score distribution due to undiagnosed cancers (which are more likely as people age). However, the higher score distributions in the younger (and exclusively female) age groups require a different explanation, which led us to examine the criteria used to include healthy donors in the Cristiano study.

The paper describing the Cristiano dataset [25] states that “[p]lasma samples from healthy individuals were obtained at the time of routine screening, including for colonoscopies or Pap smears. Individuals were considered healthy if they had no previous history of cancer and negative screening results.” This selection of healthy donors at the time of routine screening is a likely cause for the gender imbalance in the younger cohorts. Until 2018 (the year before publication of the Cristiano dataset), the American Cancer Society recommended colorectal cancer screening (often done via colonoscopy) beginning at the age of 50; this age guideline was then lowered to 45 [3]. This appears to explain the large number of healthy donors who were exactly 50 y.o. (51/241), as well as why the youngest male healthy donors are 50 y.o. Female donors could also be recruited via routine Pap smear screening, and that is almost certainly how at least some of the 23 female donors with ages < 50 y.o. were included in the study. Despite recommendations, cervical cancer screening is not widely adhered to by patients [59]. A notable exception to that can be pregnant patients, where pregnancy can be used as an opportunity to screen patients that might otherwise not be screened [60]. Therefore, the inclusion criteria used to create the Cristiano dataset may have a slight selection bias for pregnant donors, who would be screened subsequent to a positive pregnancy test. In fact, a subsequent clinical validation from many of the same authors has excluded pregnant donors from collections [30].

Fetal (or more accurately, placental) cfDNA is known to be present in blood collected from pregnant donors [61], and this is the basis for noninvasive prenatal testing [62]. Previous work has shown that the fragmentation site contexts of fetal and maternal cfDNA are distinguishable [28]. These differences between fetal and maternal cfDNA, along with the inclusion criteria used in the Cristiano study, lead us to hypothesize a cause for the elevated score distributions found in the younger, exclusively female healthy cohorts. It seems likely that fetal cfDNA is dissimilar enough to the other healthy samples that its presence can cause our similarity-based classifier to output higher (i.e., less likely to be healthy) scores. Other possible explanations could involve inflammatory diseases, such as endometriosis, that could cause elevated scores for a noncancer sample. Because the metadata available with the Cristiano dataset do not include pregnancy status or other health information for the healthy donors, we are unable to conclusively confirm or disprove these hypotheses with the data available to us.

### Tissue of origin prediction accuracy is high

When a multi-cancer screening test returns a positive result, it can often be necessary for a patient’s clinician to know where the cancer is likely to be located. Such guidance on the location is valuable to reduce the costs and side effects of imaging. Accordingly, many multi-cancer screening tests attempt to predict the tissue of origin of a cancer [19,25,29]. Given the connection between cfDNA fragmentation patterns and cellular origin [24], we also wanted to see how well the signal captured by Blackjack’s PPMs and class similarities could predict the tissue of origin.

Because the Cristiano dataset is the only one of our two that has multiple cancer types, we focused our tissue of origin examination on that dataset. We treated each of the dataset’s seven cancer types as a separate class, and used logistic regression to classify each point in a seven-dimensional class similarity space. We performed a ten-fold, 10x repeated stratified cross-validation to evaluate the accuracy of this approach (Fig. 2E, Supplementary Table S2). With each cancer type, we see at least 56% accuracy for the top prediction, with a maximum of 83% in colorectal cancer. It is also informative to see how frequently the correct tissue is in the top two predictions of the classifier [25,29]; that minimum is 82% (in gastric and lung cancer), with a maximum of 92% (in pancreatic cancer).

### No evidence of batch effects driving results

Batch effects are technical variations that are not caused by an underlying biological or scientific factor [63]. While all batches will exhibit some amount of technical variation, if batch membership correlates with sample condition (i.e., healthy/cancer), that variation may be wrongly interpreted by analysis or by ML models as biologically meaningful. We have previously shown how sequencing library batch effects related to aneuploidy caused an ML approach for ovarian cancer detection to yield overly optimistic cross-validation results [64]. Others have raised concerns about the impacts of batch effects with fragmentomics projects [65]. Because of these concerns, we investigated whether a batch effect could be an explanation for our results here. This is especially important for our work because neither of our datasets had library batching information available for the samples, and so we are unable to conclusively demonstrate if the library batches correlate with the samples’ cancer/noncancer status.

A commonly-performed step in investigating batch effects is to perform principal component analysis (PCA), and look for clusters within the samples [66]. We did such an analysis with the first two principal components (Supplementary Fig. S3), and see no obvious clusters that would indicate the healthy and cancer samples in either dataset were processed in batches separated by class. Additionally, our baseline classification approach of an Extra-Trees model using PCA provides us another avenue to investigate for the presence of batch effects. If one or more of the first 80 principal components (PCs) would discriminate well between the healthy and cancer samples, the classification model would be able to leverage those PCs to achieve high performance. In our cross-validation experiments (Fig. 2A), we see the Blackjack approach outperforms the baseline approach in both datasets. This further suggests that the variation between samples captured by PCs is not responsible for Blackjack’s performance. For all these reasons, we do not report the presence of any batch effect that impacted fragmentation site contexts in a manner that would affect the accuracy results discussed earlier.

### Library preparation differences prevent direct comparison between datasets

Although we did not find evidence that batch effects impacted fragmentation site contexts within either dataset, we do see evidence that contexts are substantially different between the two datasets. To investigate this in more detail, we restricted our analysis to the healthy samples of the two datasets. We did so because the PPMs for cancer samples can vary much more than what we see in healthy samples (Figs. 1B, 1C, 1D). On visual examination, the mean values of the PPMs’ dinucleotide frequencies are similar between the two datasets (Supplementary Figs. S4A and S4B). We do however see a noticeable difference in the percentage of fragments beginning with two C nucleotides, with the two datasets’ sample distributions not overlapping (Supplementary Fig. S4C).

For each of the 80 dinucleotide frequencies measured by our PPMs, we measured the AUC when attempting to classify between samples from the Jiang dataset and those from the Cristiano dataset. In this ROC analysis, an AUC of 0 would mean all the Cristiano healthy samples had a dinucleotide frequency higher than the corresponding frequency in all the Jiang healthy samples. An AUC of 1 would also imply perfect separation, but with frequencies ordered oppositely. AUCs closer to 0.5 indicate that the dinucleotide frequency does not distinguish well between the two datasets’ healthy samples. Eight of the 80 dinucleotide frequencies provide perfect classification (Supplementary Fig. S4D), and 59 of 80 AUCs are either less than 0.2 or greater than 0.8. This means that many of the frequencies that constitute our PPMs are so different between the two datasets that complex ML classifiers are not needed to determine to which dataset a given sample belongs.

The differences between the two datasets’ healthy samples are also clearly seen in the first principal component (PC1), which completely separates the two datasets (Supplementary Fig. S4E). The leading CC percentage is highly anticorrelated with PC1, with a Pearson correlation coefficient of -0.99. Additionally, the distances between healthy samples within a dataset are far less than the distances between healthy samples across datasets (Supplementary Fig. S4F). The distribution of HCD values is also considerably different between the two datasets’ healthy samples (Jiang mean = 0.0073, s.d. = 0.0032; Cristiano mean = 0.0101, s.d. = 0.00329). This increased variability (absolute deviation from the centroid) of healthy samples in the Cristiano dataset can also be seen in Fig. 1C.

We hypothesize that these differences between the two datasets’ PPMs are the result of different forms of ligation bias caused by different library preparation methods and reagents. Ligation bias can alter observed base composition of positions both upstream and downstream of sequenced fragment ends [67], and different library preparation kits can result in different observed base compositions [68]. The Jiang dataset was generated using the Kapa Library Preparation Kit [35], while the Cristiano dataset was generated using the NEBNext DNA Library Prep Kit [25], along with some modifications. Even if ligation bias is not the root cause of the observed context differences discussed here, the differences frustrate cross-dataset comparisons or applications of ML across the datasets.

### Performance is stable at very low sequencing depth

The columns of our PPMs are empirical dinucleotide PMFs in their own right, with only 15 free parameters per column under the unit-sum constraint. Because the positional independence assumption of the PPM allows each column to be estimated separately from the same set of observed hexamers, and because each fragment end contributes an observation to every column, we expected that the PPMs would converge to stable values even with relatively little sequencing data. To test this, we used the hexamers for the Cristiano dataset to create a hexamer PMF for each individual. These hexamer PMFs then let us generate a set of new hexamers drawn from the same observed distributions as were seen in the original dataset, but at different simulated sequencing depths. In this way, we could simulate how well the Blackjack algorithm would perform when, for example, only 1 million fragments per sample were sequenced. Comparing the retrospective results (Fig. 1F) against these simulated results allows us to understand the relationship between sequencing depth and Blackjack’s performance.

For each depth we examined, we simulated the Cristiano dataset 10 times; we show the results of these simulations in Supplementary Fig. 5. Using 4 million simulated fragments per sample, we see a mean AUC of 0.993 and a mean sensitivity (at 98% specificity) of 90.3%. This performance is close to the full-dataset retrospective AUC of 0.994 and sensitivity of 91.5%. We also see similar although lesser performance at 1 million simulated fragments, with a mean AUC of 0.991 and mean sensitivity of 87.6%. Lower amounts of simulated sequencing suggest that performance would begin to degrade as depth decreases below 1 million fragments. Whether lower depths are viable for a screening test would depend on the required specificity and sensitivity. When measured at 70% specificity, as might be used when testing a symptomatic or high-risk population [30,69], sensitivity remains above 99% at 250,000 simulated fragments.

Using 100 bp paired-end reads, 4 million fragments of human WGS data corresponds to an effective depth of 0.24x. In contrast, approximately 3x was used in the clinical validation of a commercially-available fragmentomics-based cancer detection test [30]. These results suggest a Blackjack-based test would require substantially less sequencing.

## Discussion

Blackjack’s fragmentomics approach focuses on modelling fragmentation sites through their sequence context (in PPMs), and modelling the healthy and cancer populations via a combination of centroids and covariance matrices. By representing an individual’s cfDNA fragmentation profile as a PPM, Blackjack can achieve high accuracy for cancer detection while requiring only small amounts of sequencing (and therefore, a small amount of input cfDNA). Blackjack also achieves high accuracy on early-stage cancers, a particularly important subset given the improvements to mortality that are seen when cancer is detected early [1]. With further work, the high sensitivity of Blackjack on early-stage cancers could be used to improve or complement existing early detection and monitoring assays.

While others have examined sequence contexts for fragmentomics, we have not seen AUC results approaching the minimum of 0.993 we see in our datasets (Fig. 2A). The motif diversity score (MDS), an entropy-based measure, has been employed to distinguish between cancer and noncancer samples [28]. MDS calculation relies on the 256 frequencies of tetramers found at the internal ends of fragments. The MDS is a single number computed for a sample (effectively reducing dimensionality from 256 to 1), without reference to other samples; after calculation, a sample’s MDS value would then be compared against that of other known samples. In contrast, Blackjack’s supervised dimensionality reduction uses comparisons to other samples, and may be retaining more valuable information as a result. Another method, GALYFRE, examines some mononucleotide frequencies around the fragmentation sites along with other features [31]. These features are then passed directly into a random forest classifier. Given that our baseline classifier directly uses PCA-transformed dinucleotide frequencies, and achieves lower performance than Blackjack (Fig. 2A), it appears that the similarity-based dimensionality reductions are providing value beyond that of the frequencies alone. Further work would be needed to directly compare Blackjack against these other methods, or to determine exactly how a method is outperforming another.

Our results indicate that sequence context around the observed fragmentation sites can be a powerful tool for distinguishing between healthy and cancer samples. However, we also see important implications in these results on the laboratory methods used to analyze cfDNA. Elements of double-stranded DNA library preparation, especially end repair [70], can alter the observed fragment ends of cfDNA. Recent work [71] has shown that single-stranded library preparation kits [72] may be valuable in obtaining a more accurate representation of cfDNA fragment ends. Regardless of whether double- or single-stranded library preparation is used, it will be important to ensure that future work holds the laboratory protocols and preparation kits constant between training, validation, and any production use of methods that rely on analyzing the sites of fragmentation.

The results we obtained from previously-published datasets also illustrate the value of detailed metadata and methodology when using ML approaches in diagnostics and screening. The presence of age and sex information for the healthy donors of the Cristiano dataset allowed us to see the elevated scores within the younger female cohort, and our investigation then led to our hypothesis that those scores were likely due to pregnant donors who were recruited into the healthy cohort. This hypothesis would not have been formed had the metadata not been available, and had the recruitment criteria not been stated in the original paper. Conversely, had library batch data been available for our datasets, we would have been able to do more rigorous analysis on the possibility of a batch effect driving our results. As we move forward with future investigations into the value of fragmentomics with our own sequencing data, we will endeavor to provide such metadata so that it can enable downstream analyses by others.

With regard to future work, it is important to note the limitations of this study. Our results here were only cross-validated, and must necessarily be limited to the datasets used. It is also difficult, if not impossible, to know how a classification method would perform on incipient cancers when the validation is not performed in a large asymptomatic population, with clinical followup to confirm results. In future validation work, we will need to ensure that training and validation data are generated with the same laboratory protocols. In designing new training and validation laboratory experiments, we will also need to track library batch information as well as provide a temporal separation between the processing of the training samples and the validation samples. Such a separation would provide assurance that the performance on the validation data was not influenced by data leakage [73] that would yield overly optimistic results.

The high cross-validated performance of Blackjack on early-stage cancers (Fig. 2B) suggests that cfDNA fragmentation patterns, and specifically the sites of fragmentation, can be of great value in early cancer detection and in detection of low levels of recurrent disease. Yet the results we see in comparing healthy samples across datasets show that development of fragmentomics solutions may require the construction of a new dataset for a specific test. It is also likely that certain WGS protocols will provide better ability to distinguish between healthy and cancer samples when examining fragmentation site context data. Further development of fragmentomics-based cancer tests may benefit from a focus on minimizing variability of observed fragmentation site signals, and conclusive demonstration of robustness to batch effects.

When designing Blackjack, we wanted to minimize the opportunity for our models to overfit. We did this by keeping the feature space of our LR classification models small, and using hold-out calculations for class centroids and covariance matrices when computing distances during model fitting. These steps appear to have worked to make the retrospective AUCs for our distance- and similarity-based methods (Figs. 1C, 1D, 1F) very similar to the AUC results we see in the 10-fold CV (Fig. 2A) and even the 2-fold CV (Supplementary Fig. S2). While future work is still necessary to conclusively demonstrate that our approach can work well on new data, the low differences between the retrospective and CV results are promising.

Finally, the two features of Blackjack’s LR model are quantifying answers to simple questions: how similar is an individual’s fragmentation pattern to that of (a) other known healthy samples, and (b) other known cancer samples? This focus on a low number of informative and interpretable features is valuable for training in situations where the total number of training data points (i.e., donor samples) is small, as is the case with the datasets we used here. Having more features than would be supported by the training dataset can lead to overfit ML models that may not generalize to new data [74,75], even when regularization penalties are employed [64]. When each training data point represents an individual who has donated plasma, acquiring more training data can be a costly endeavor; far more costly is the effect of a failed validation or clinical trial when an overfit ML model does not perform to expectations. Therefore, in the development of ML approaches for diagnostics, we will continue to seek parsimonious solutions that will be more likely to reproduce their cross-validation results on new data.

## Supplementary Figures

**Supplementary Figure S1.**
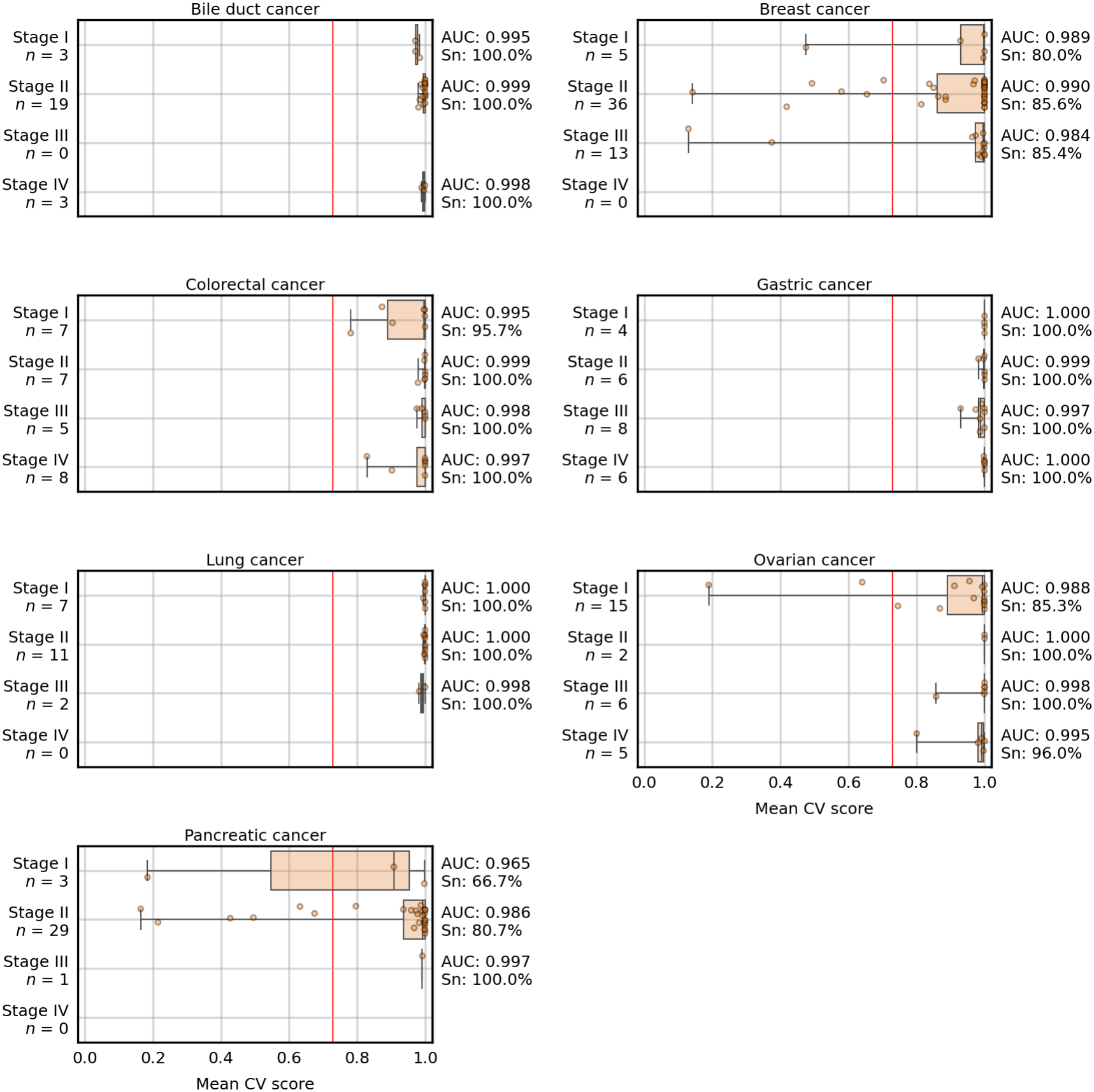
Cross-validation results for the Cristiano dataset cancer samples. Results for the Blackjack approach are stratified by cancer type and stage. AUC and Sn at 98% Sp are shown for each approach, and the 98% Sp score threshold is shown as a vertical red line.

**Supplementary Figure S2.**
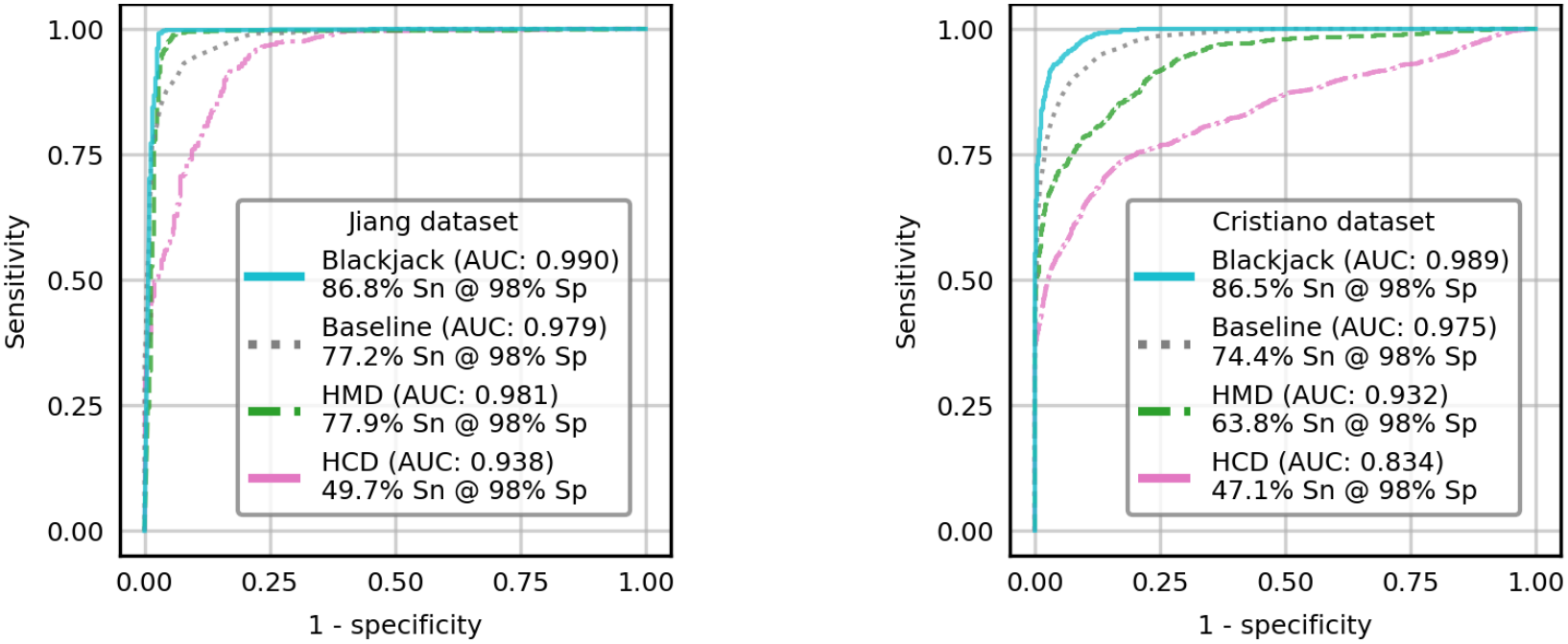
Two-fold cross-validation results for the two datasets. To evaluate the robustness of the methods we examined, we also performed a two-fold cross-validation experiment with each dataset.

**Supplementary Figure S3.**
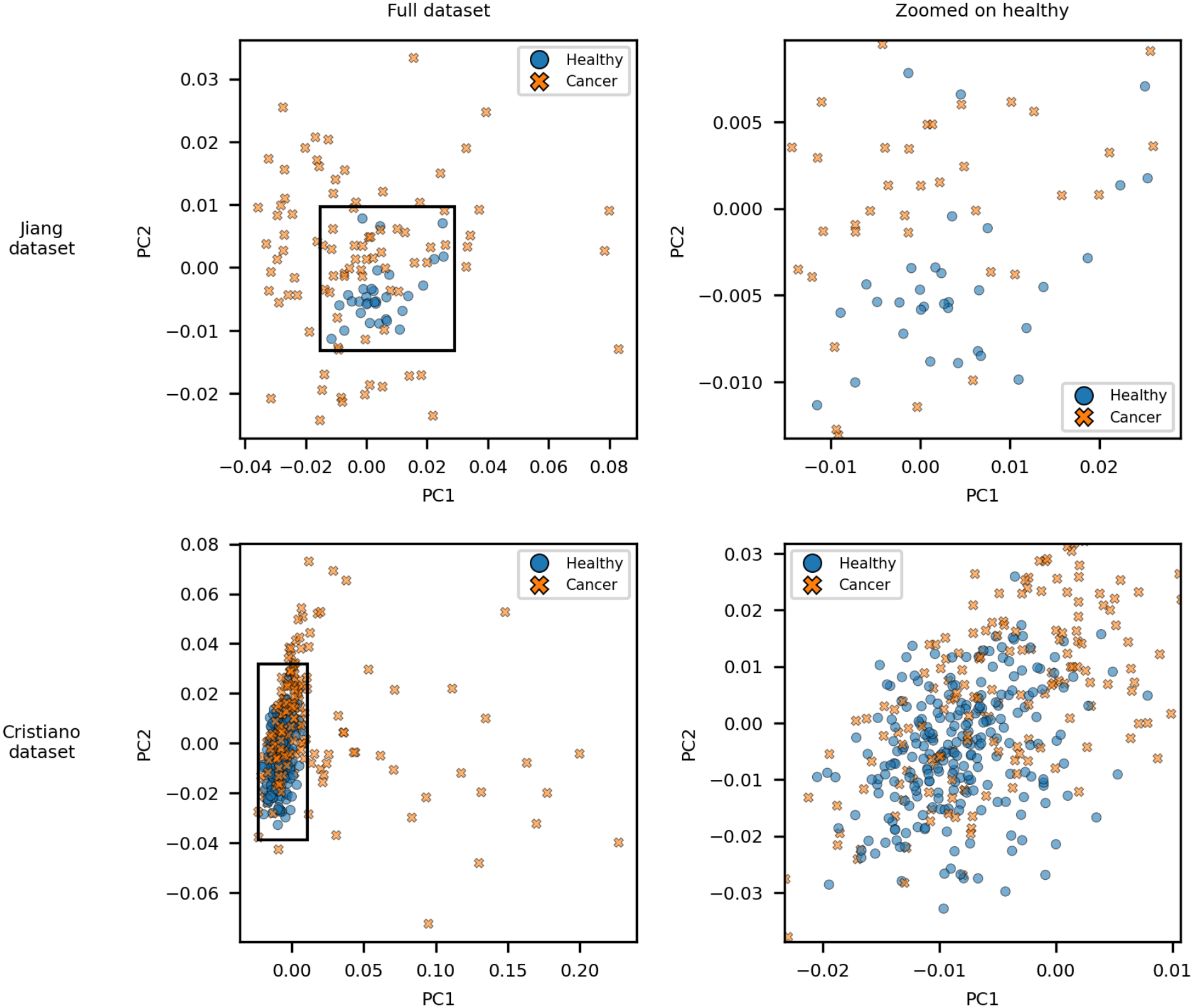
Two-dimensional PCA of PPM data from the two datasets. A principal component analysis of both datasets shows no evidence of clusters that would indicate batch effects influenced our classification results.

**Supplementary Figure S4.**
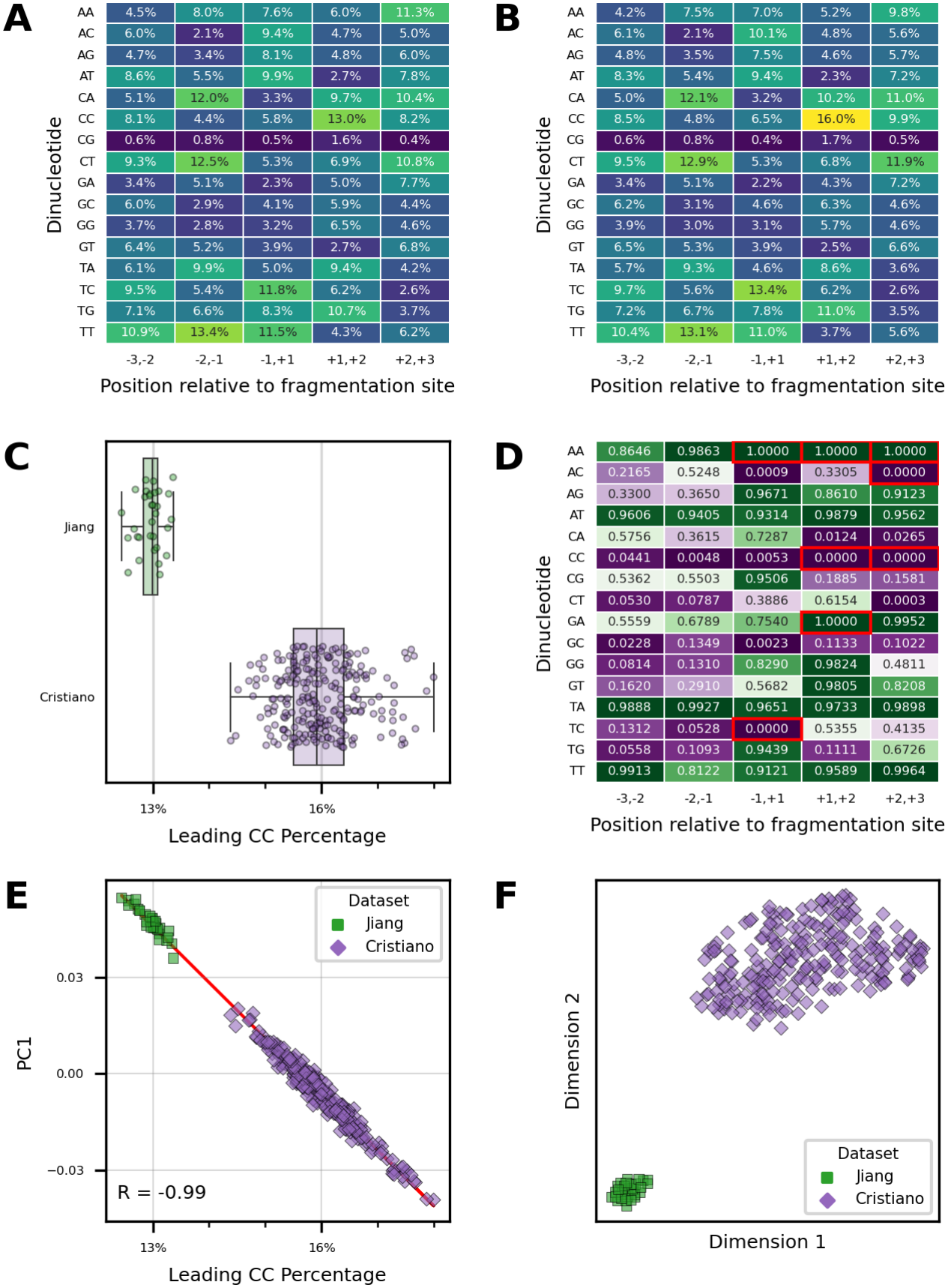
Differences between healthy samples of the two datasets. **(A)** Mean PPM values for healthy samples in the Jiang dataset. **(B)** Mean PPM values for healthy samples in the Cristiano dataset. **(C)** The percentage of each healthy sample’s fragments that begin with two C nucleotides, separated by dataset. **(D)** Receiver operating characteristic analysis of dataset prediction, with area under the curve (AUC) shown for each dinucleotide frequency feature. An AUC of 1 corresponds to the feature being higher in all Jiang healthy samples than in all Cristiano healthy samples. Eight dinucleotide frequencies that are perfect classifiers are outlined in red. **(E)** Principal component analysis of the healthy samples shows the first principal component (PC1) provides complete separation between the two datasets, and is highly anticorrelated with the percentage of sequenced fragments that begin with a pair of C nucleotides. The linear regression line is shown in red, with Pearson correlation coefficient (R) reported in the figure. **(F)** t-SNE visualization of the two datasets also shows strong separation between the two datasets.

**Supplementary Figure S5.**
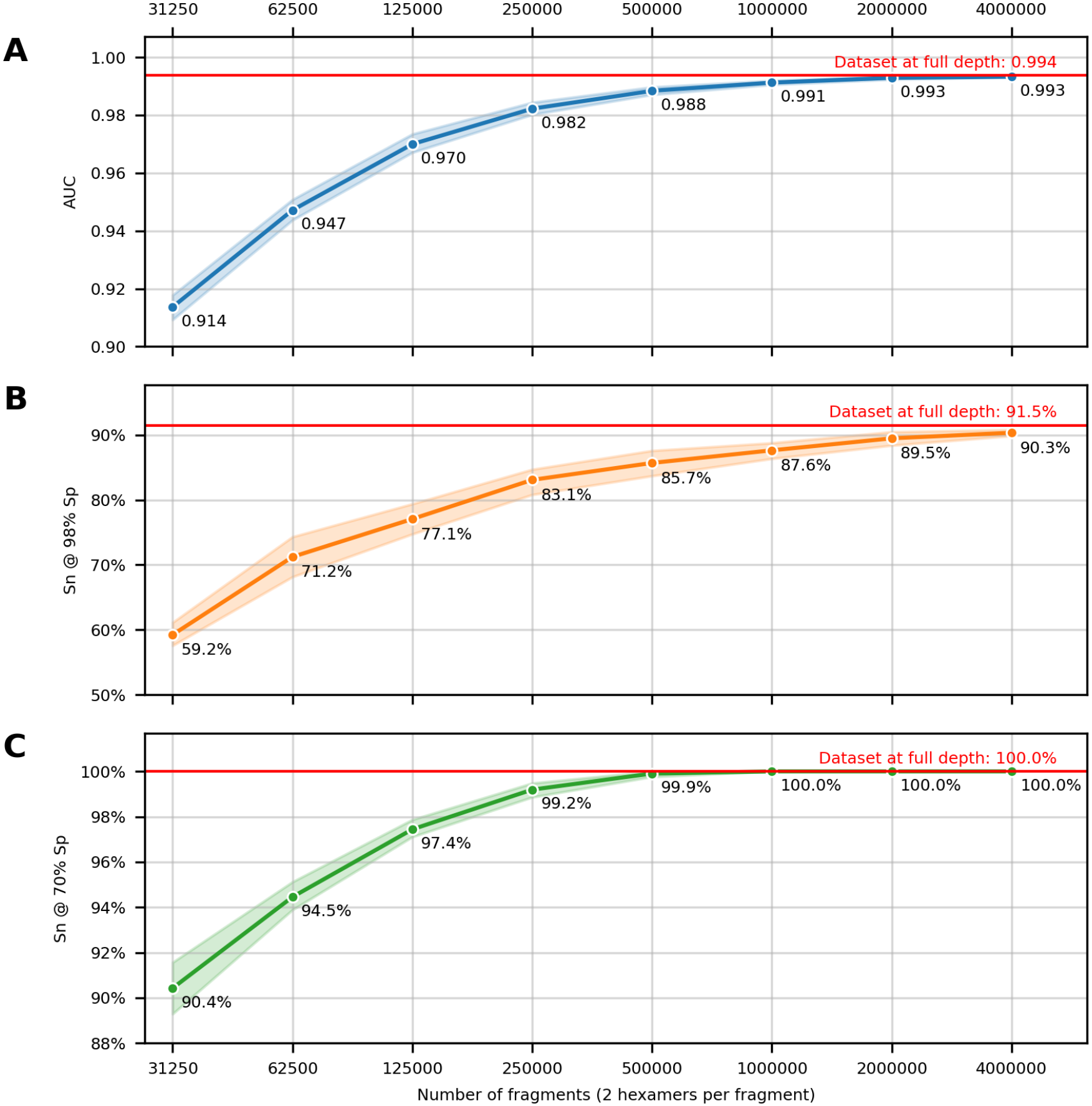
Accuracy at different simulated sequencing depths. **(A)** Receiver operating characteristic area under the curve (AUC) of Cristiano dataset prediction. **(B)** Sensitivity (Sn) at 98% specificity (Sp). **(C)** Sensitivity at 70% specificity. A red horizontal line represents the corresponding accuracy statistic for the full Cristiano dataset in each panel. Each curve is the mean value with a 95% confidence interval indicated by the lighter surrounding regions; all x-axes are on a logarithmic scale.

## Supplementary Tables

**Supplementary Table S1.**
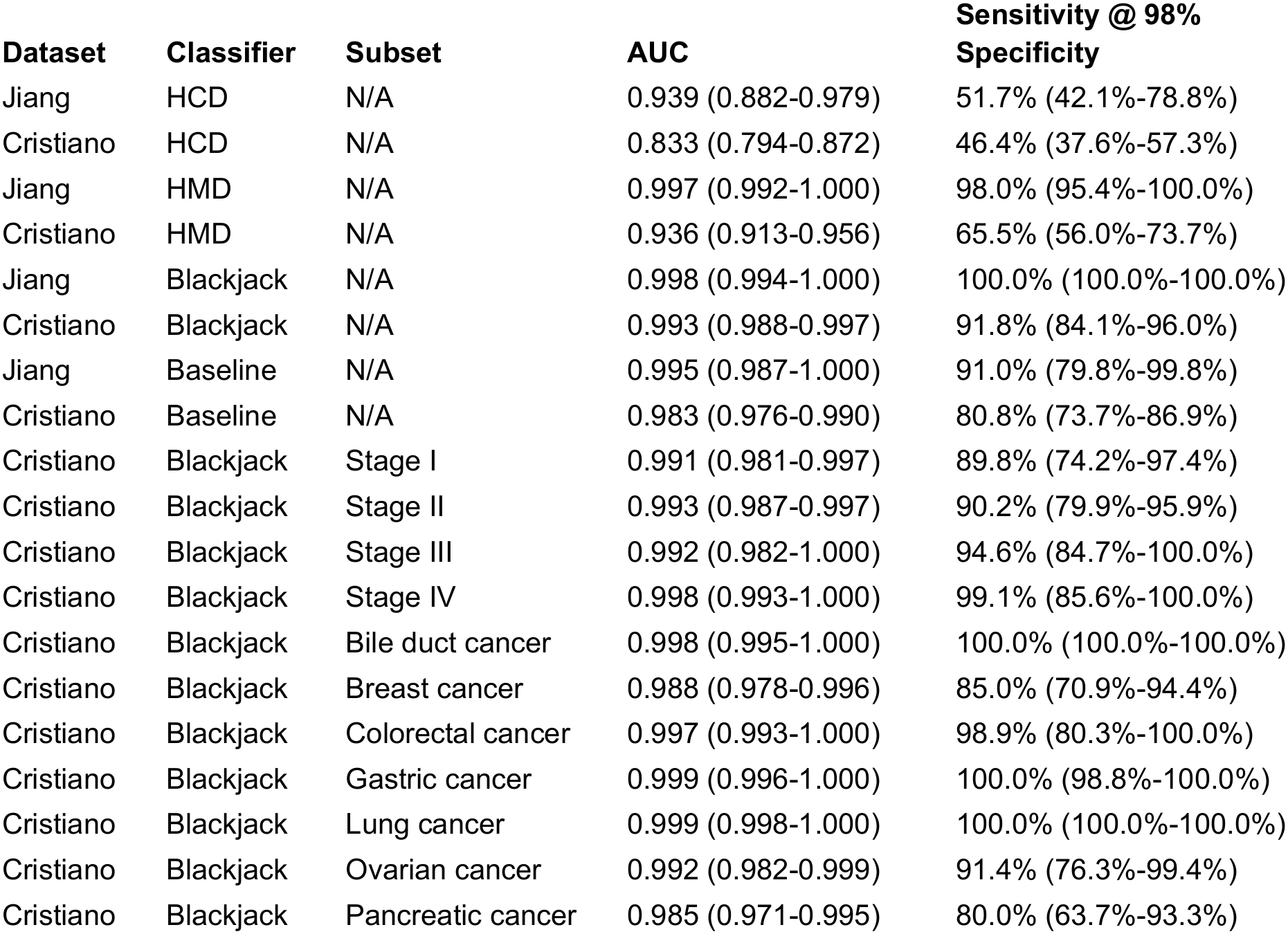
Point estimates and 95% confidence intervals for cancer detection CV experiments.

**Supplementary Table S2.** Point estimates and 95% confidence intervals for tissue of origin CV experiments.

| **Cancer type** | **Top Prediction Accuracy** | **Top 2 Predictions Accuracy** |
|---|---|---|
| Bile duct cancer | 74% (58%-88%) | 86% (72%-97%) |
| Breast cancer | 81% (72%-88%) | 91% (84%-97%) |
| Colorectal cancer | 83% (69%-94%) | 89% (77%-98%) |
| Gastric cancer | 78% (61%-91%) | 82% (67%-95%) |
| Lung cancer | 56% (34%-76%) | 82% (69%-92%) |
| Ovarian cancer | 66% (51%-82%) | 83% (71%-93%) |
| Pancreatic cancer | 83% (72%-93%) | 92% (84%-98%) |

## Data Availability

The source processed sequencing data were obtained from FinaleDB (http://finaledb.research.cchmc.org/). Software to produce the results of this manuscript is available at http://dx.doi.org/10.5281/zenodo.20768356.

## References

1. Crosby D, Bhatia S, Brindle KM, Coussens LM, Dive C, Emberton M, et al. Early detection of cancer. Science. 2022;375: eaay9040. doi:10.1126/science.aay9040

2. Siegel RL, Kratzer TB, Wagle NS, Sung H, Jemal A. Cancer statistics, 2026. CA Cancer J Clin. 2026;76: e70043. doi:10.3322/caac.70043

3. Wolf AMD, Fontham ETH, Church TR, Flowers CR, Guerra CE, LaMonte SJ, et al. Colorectal cancer screening for average-risk adults: 2018 guideline update from the American Cancer Society: ACS Colorectal Cancer Screening Guideline. CA Cancer J Clin. 2018;68: 250–281. doi:10.3322/caac.21457

4. Oeffinger KC, Fontham ETH, Etzioni R, Herzig A, Michaelson JS, Shih Y-CT, et al. Breast cancer screening for women at average risk: 2015 guideline update from the American Cancer Society. JAMA. 2015;314: 1599–1614. doi:10.1001/jama.2015.12783

5. Perkins RB, Wolf AMD, Church TR, Elkin EB, Skates SJ, Etzioni RD, et al. Self-collected vaginal specimens for human papillomavirus testing and guidance on screening exit: An update to the American Cancer Society cervical cancer screening guideline. CA Cancer J Clin. 2026;76: e70041. doi:10.3322/caac.70041

6. Goossens M, Tran TN. The effect on attendance of the requirement to confirm a pre-scheduled appointment in a population-based mammography screening programme. Eur J Public Health. 2025;35: 353–358. doi:10.1093/eurpub/ckaf028

7. Denberg TD, Melhado TV, Coombes JM, Beaty BL, Berman K, Byers TE, et al. Predictors of nonadherence to screening colonoscopy. J Gen Intern Med. 2005;20: 989–995. doi:10.1111/j.1525-1497.2005.00164.x

8. Hoyo C, Yarnall KSH, Skinner CS, Moorman PG, Sellers D, Reid L. Pain predicts non-adherence to pap smear screening among middle-aged African American women. Prev Med. 2005;41: 439–445. doi:10.1016/j.ypmed.2004.11.021

9. US Preventive Services Task Force, Nicholson WK, Silverstein M, Wong JB, Barry MJ, Chelmow D, et al. Screening for breast cancer: US preventive services task force recommendation statement. JAMA. 2024;331: 1918–1930. doi:10.1001/jama.2024.5534

10. US Preventive Services Task Force, Davidson KW, Barry MJ, Mangione CM, Cabana M, Caughey AB, et al. Screening for colorectal cancer: US Preventive Services Task Force recommendation statement. JAMA. 2021;325: 1965–1977. doi:10.1001/jama.2021.6238

11. US Preventive Services Task Force, Krist AH, Davidson KW, Mangione CM, Barry MJ, Cabana M, et al. Screening for lung cancer: US Preventive Services Task Force recommendation statement. JAMA. 2021;325: 962–970. doi:10.1001/jama.2021.1117

12. US Preventive Services Task Force, Curry SJ, Krist AH, Owens DK, Barry MJ, Caughey AB, et al. Screening for cervical cancer: US Preventive Services Task Force recommendation statement. JAMA. 2018;320: 674–686. doi:10.1001/jama.2018.10897

13. Connal S, Cameron JM, Sala A, Brennan PM, Palmer DS, Palmer JD, et al. Liquid biopsies: the future of cancer early detection. J Transl Med. 2023;21: 118. doi:10.1186/s12967-023-03960-8

14. Russo A, Incorvaia L, Del Re M, Malapelle U, Capoluongo E, Gristina V, et al. The molecular profiling of solid tumors by liquid biopsy: a position paper of the AIOM-SIAPEC-IAP-SIBioC-SIC-SIF Italian Scientific Societies. ESMO Open. 2021;6: 100164. doi:10.1016/j.esmoop.2021.100164

15. Honoré N, Galot R, van Marcke C, Limaye N, Machiels J-P. Liquid biopsy to detect minimal residual disease: Methodology and impact. Cancers (Basel). 2021;13: 5364. doi:10.3390/cancers13215364

16. Bast RC Jr, Klug TL, St John E, Jenison E, Niloff JM, Lazarus H, et al. A radioimmunoassay using a monoclonal antibody to monitor the course of epithelial ovarian cancer. N Engl J Med. 1983;309: 883–887. doi:10.1056/NEJM198310133091503

17. Mathieu KB, Bedi DG, Thrower SL, Qayyum A, Bast RC Jr. Screening for ovarian cancer: imaging challenges and opportunities for improvement. Ultrasound Obstet Gynecol. 2018;51: 293–303. doi:10.1002/uog.17557

18. Buys SS, Partridge E, Greene MH, Prorok PC, Reding D, Riley TL, et al. Ovarian cancer screening in the Prostate, Lung, Colorectal and Ovarian (PLCO) cancer screening trial: findings from the initial screen of a randomized trial. Am J Obstet Gynecol. 2005;193: 1630–1639. doi:10.1016/j.ajog.2005.05.005

19. Alexander GE, Lin W, Ortega FE, Ramaiah M, Jung B, Ji L, et al. Analytical validation of a multi-cancer early detection test with cancer signal origin using a cell-free DNA-based targeted methylation assay. PLoS One. 2023;18: e0283001. doi:10.1371/journal.pone.0283001

20. Lennon AM, Buchanan AH, Kinde I, Warren A, Honushefsky A, Cohain AT, et al. Feasibility of blood testing combined with PET-CT to screen for cancer and guide intervention. Science. 2020;369: eabb9601. doi:10.1126/science.abb9601

21. Douville C, Cohen JD, Ptak J, Popoli M, Schaefer J, Silliman N, et al. Assessing aneuploidy with repetitive element sequencing. Proc Natl Acad Sci U S A. 2020;117: 4858–4863. doi:10.1073/pnas.1910041117

22. Imai M, Nakamura Y, Yoshino T. Transforming cancer screening: the potential of multi-cancer early detection (MCED) technologies. Int J Clin Oncol. 2025;30: 180–193. doi:10.1007/s10147-025-02694-5

23. Tsui WHA, Jiang P, Lo YMD. Cell-free DNA fragmentomics in cancer. Cancer Cell. 2025;43: 1792–1814. doi:10.1016/j.ccell.2025.09.006

24. Snyder MW, Kircher M, Hill AJ, Daza RM, Shendure J. Cell-free DNA comprises an in vivo nucleosome footprint that informs its tissues-of-origin. Cell. 2016;164: 57–68. doi:10.1016/j.cell.2015.11.050

25. Cristiano S, Leal A, Phallen J, Fiksel J, Adleff V, Bruhm DC, et al. Genome-wide cell-free DNA fragmentation in patients with cancer. Nature. 2019;570: 385–389. doi:10.1038/s41586-019-1272-6

26. Mouliere F, Chandrananda D, Piskorz AM, Moore EK, Morris J, Ahlborn LB, et al. Enhanced detection of circulating tumor DNA by fragment size analysis. Sci Transl Med. 2018;10: eaat4921. doi:10.1126/scitranslmed.aat4921

27. Underhill HR, Kitzman JO, Hellwig S, Welker NC, Daza R, Baker DN, et al. Fragment length of circulating tumor DNA. PLoS Genet. 2016;12: e1006162. doi:10.1371/journal.pgen.1006162

28. Jiang P, Sun K, Peng W, Cheng SH, Ni M, Yeung PC, et al. Plasma DNA end-motif profiling as a fragmentomic marker in cancer, pregnancy, and transplantation. Cancer Discov. 2020;10: 664–673. doi:10.1158/2159-8290.CD-19-0622

29. Zhou X, Zheng H, Fu H, Dillehay McKillip KL, Pinney SM, Liu Y. CRAG: de novo characterization of cell-free DNA fragmentation hotspots in plasma whole-genome sequencing. Genome Med. 2022;14: 138. doi:10.1186/s13073-022-01141-8

30. Mazzone PJ, Bach PB, Carey J, Schonewolf CA, Bognar K, Ahluwalia MS, et al. Clinical validation of a cell-free DNA fragmentome assay for augmentation of lung cancer early detection. Cancer Discov. 2024;14: 2224–2242. doi:10.1158/2159-8290.CD-24-0519

31. Budhraja KK, McDonald BR, Stephens MD, Contente-Cuomo T, Markus H, Farooq M, et al. Genome-wide analysis of aberrant position and sequence of plasma DNA fragment ends in patients with cancer. Sci Transl Med. 2023;15: eabm6863. doi:10.1126/scitranslmed.abm6863

32. Fostier J. BLAMM: BLAS-based algorithm for finding position weight matrix occurrences in DNA sequences on CPUs and GPUs. BMC Bioinformatics. 2020;21: 81. doi:10.1186/s12859-020-3348-6

33. Zheng H, Zhu MS, Liu Y. FinaleDB: a browser and database of cell-free DNA fragmentation patterns. Bioinformatics. 2021;37: 2502–2503. doi:10.1093/bioinformatics/btaa999

34. Stormo GD. DNA binding sites: representation and discovery. Bioinformatics. 2000;16: 16–23. doi:10.1093/bioinformatics/16.1.16

35. Jiang P, Chan CWM, Chan KCA, Cheng SH, Wong J, Wong VW-S, et al. Lengthening and shortening of plasma DNA in hepatocellular carcinoma patients. Proc Natl Acad Sci U S A. 2015;112: E1317–25. doi:10.1073/pnas.1500076112

36. Valentine AP, Sambridge M. Optimal regularization for a class of linear inverse problem. Geophys J Int. 2018;215: 1003–1021. doi:10.1093/gji/ggy303

37. Rousseeuw PJ, van Zomeren BC. Unmasking multivariate outliers and leverage points. J Am Stat Assoc. 1990;85: 633. doi:10.2307/2289995

38. Hastie T. Ridge regularization: An essential concept in data science. Technometrics. 2020;62: 426–433. doi:10.1080/00401706.2020.1791959

39. Geurts P, Ernst D, Wehenkel L. Extremely randomized trees. Mach Learn. 2006;63: 3–42. doi:10.1007/s10994-006-6226-1

40. Pedregosa F, Varoquaux G, Gramfort A, Michel V, Thirion B, Grisel O, et al. Scikit-learn: Machine Learning in Python. arXiv [cs.LG]. 2018. doi:10.48550/arXiv.1201.0490

41. Hunter JD. Matplotlib: A 2D Graphics Environment. Comput Sci Eng. 2007;9: 90–95. doi:10.1109/mcse.2007.55

42. Waskom M. seaborn: statistical data visualization. J Open Source Softw. 2021;6: 3021. doi:10.21105/joss.03021

43. McKinney W. Data Structures for Statistical Computing in Python. Proceedings of the Python in Science Conference. SciPy; 2010. pp. 56–61. doi:10.25080/majora-92bf1922-00a

44. Harris CR, Millman KJ, van der Walt SJ, Gommers R, Virtanen P, Cournapeau D, et al. Array programming with NumPy. Nature. 2020;585: 357–362. doi:10.1038/s41586-020-2649-2

45. Virtanen P, Gommers R, Oliphant TE, Haberland M, Reddy T, Cournapeau D, et al. SciPy 1.0: fundamental algorithms for scientific computing in Python. Nat Methods. 2020;17: 261–272. doi:10.1038/s41592-019-0686-2

46. Quinlan AR, Hall IM. BEDTools: a flexible suite of utilities for comparing genomic features. Bioinformatics. 2010;26: 841–842. doi:10.1093/bioinformatics/btq033

47. Li H, Handsaker B, Wysoker A, Fennell T, Ruan J, Homer N, et al. The Sequence Alignment/Map format and SAMtools. Bioinformatics. 2009;25: 2078–2079. doi:10.1093/bioinformatics/btp352

48. Wood DE. Software and data for Blackjack method paper. 2026. doi:10.5281/zenodo.20768356

49. Korhonen JH, Palin K, Taipale J, Ukkonen E. Fast motif matching revisited: high-order PWMs, SNPs and indels. Bioinformatics. 2017;33: 514–521. doi:10.1093/bioinformatics/btw683

50. Lander ES, Linton LM, Birren B, Nusbaum C, Zody MC, Baldwin J, et al. Initial sequencing and analysis of the human genome. Nature. 2001;409: 860–921. doi:10.1038/35057062

51. Mathelier A, Wasserman WW. The next generation of transcription factor binding site prediction. PLoS Comput Biol. 2013;9: e1003214. doi:10.1371/journal.pcbi.1003214

52. Endres DM, Schindelin JE. A new metric for probability distributions. IEEE Trans Inf Theory. 2003;49: 1858–1860. doi:10.1109/tit.2003.813506

53. Van der Maaten L, Hinton G. Visualizing data using t-SNE. J Mach Learn Res. 2008;9: 2579−2605. Available: https://www.jmlr.org/papers/volume9/vandermaaten08a/vandermaaten08a.pdf

54. Fan J, Upadhye S, Worster A. Understanding receiver operating characteristic (ROC) curves. CJEM. 2006;8: 19–20. doi:10.1017/s1481803500013336

55. Mahalanobis PC. On the generalised distance in statistics. Proc Natl Inst Sci India. 1936;2: 49–55.

56. De Santis E, Martino A, Rizzi A. On component-wise dissimilarity measures and metric properties in pattern recognition. PeerJ Comput Sci. 2022;8: e1106. doi:10.7717/peerj-cs.1106

57. Pinsky PF. Principles of cancer screening. Surg Clin North Am. 2015;95: 953–966. doi:10.1016/j.suc.2015.05.009

58. Arisi MF, Dotan E, Fernandez SV. Circulating tumor DNA in precision oncology and its applications in colorectal cancer. Int J Mol Sci. 2022;23: 4441. doi:10.3390/ijms23084441

59. Shin MB, Axeen S, Cole AM, Guo XM, Islam JY, Ko LK, et al. Nonadherence to cervical cancer screening guidelines in commercially insured US adults, 2013-2021. JAMA Netw Open. 2025;8: e2548512. doi:10.1001/jamanetworkopen.2025.48512

60. Kumari S. Screening for cervical cancer in pregnancy. Oncol Rev. 2023;17: 11429. doi:10.3389/or.2023.11429

61. Dugoff L, Barberio A, Whittaker PG, Schwartz N, Sehdev H, Bastek JA. Cell-free DNA fetal fraction and preterm birth. Am J Obstet Gynecol. 2016;215: 231.e1–7. doi:10.1016/j.ajog.2016.02.009

62. Taglauer ES, Wilkins-Haug L, Bianchi DW. Review: cell-free fetal DNA in the maternal circulation as an indication of placental health and disease. Placenta. 2014;35 Suppl: S64–8. doi:10.1016/j.placenta.2013.11.014

63. Leek JT, Scharpf RB, Bravo HC, Simcha D, Langmead B, Johnson WE, et al. Tackling the widespread and critical impact of batch effects in high-throughput data. Nat Rev Genet. 2010;11: 733–739. doi:10.1038/nrg2825

64. Wood DE, Roy J, Ballew BJ. Simpler predictive models provide higher accuracy for ovarian cancer detection. PeerJ. 2025;13: e20525. doi:10.7717/peerj.20525

65. Liu Y. At the dawn: cell-free DNA fragmentomics and gene regulation. Br J Cancer. 2022;126: 379–390. doi:10.1038/s41416-021-01635-z

66. Reese SE, Archer KJ, Therneau TM, Atkinson EJ, Vachon CM, de Andrade M, et al. A new statistic for identifying batch effects in high-throughput genomic data that uses guided principal component analysis. Bioinformatics. 2013;29: 2877–2883. doi:10.1093/bioinformatics/btt480

67. Seguin-Orlando A, Schubert M, Clary J, Stagegaard J, Alberdi MT, Prado JL, et al. Ligation bias in illumina next-generation DNA libraries: implications for sequencing ancient genomes. PLoS One. 2013;8: e78575. doi:10.1371/journal.pone.0078575

68. Baldrich P, Tamim S, Mathioni S, Meyers B. Ligation bias is a major contributor to nonstoichiometric abundances of secondary siRNAs and impacts analyses of microRNAs. bioRxiv. bioRxiv; 2020. doi:10.1101/2020.09.14.296616

69. Law RA, Giles BM, Culp-Hill R, Radnaa E, Goldberg M, Nichols CM, et al. Clinical evaluation of a multi-omic diagnostic model for early-stage ovarian cancer detection. Diagnostics (Basel). 2025;15: 2225. doi:10.3390/diagnostics15172225

70. Groth TE, Mishin AA, Rao V, Tibet R, Troll CJ. End-repair causes methylation underestimation in cell-free DNA sequencing libraries. J Liq Biopsy. 2026;11: 100455. doi:10.1016/j.jlb.2026.100455

71. Jiang P, Ma M-JL, Qiao R, Shi Y, Liu J, Zhou Q, et al. Holistic determination of ends of cfDNA molecules. Cell Genom. 2026;6: 101142. doi:10.1016/j.xgen.2026.101142

72. Troll CJ, Kapp J, Rao V, Harkins KM, Cole C, Naughton C, et al. A ligation-based single-stranded library preparation method to analyze cell-free DNA and synthetic oligos. BMC Genomics. 2019;20: 1023. doi:10.1186/s12864-019-6355-0

73. Kaufman S, Rosset S, Perlich C. Leakage in data mining: formulation, detection, and avoidance. Proceedings of the 17th ACM SIGKDD international conference on Knowledge discovery and data mining. New York, NY, USA: ACM; 2011. doi:10.1145/2020408.2020496

74. Stoltzfus JC. Logistic regression: a brief primer: Logistic regression: A brief primer. Acad Emerg Med. 2011;18: 1099–1104. doi:10.1111/j.1553-2712.2011.01185.x

75. Divasón J, Ceniceros JF, Sanz-Garcia A, Pernia-Espinoza A, Martinez-de-Pison FJ. PSO-PARSIMONY: A method for finding parsimonious and accurate machine learning models with particle swarm optimization. Application for predicting force–displacement curves in T-stub steel connections. Neurocomputing. 2023;548: 126414. doi:10.1016/j.neucom.2023.126414

